# How did COVID-19 measures impact sexual behaviour and access to HIV/STI services in Panama? Results from a national cross-sectional online survey

**DOI:** 10.1101/2021.02.03.21251095

**Authors:** Amanda Gabster, Jennifer Toller Erausquin, Kristien Michielsen, Philippe Mayaud, Juan Miguel Pascale, Carles Pericas Escalé, Michael Marks, Jennifer Katz, Gonzalo Cabezas Talavero, Marilu de Argote, Anet Murillo, Joseph D. Tucker

**Affiliations:** Instituto Conmemorativo Gorgas de Estudios de la Salud, Ave. Justo Arosemena y Calle 36, Panamá, Panamá; Instituto Conmemorativo Gorgas de Estudios de la Salud, Panama; London School of Hygiene and Tropical Medicine, UK, Faculty of Infectious and Tropical Diseases; University of North Carolina Greensboro, USA, School of Health and Human Sciences; Universiteit Gent, Belgium, Faculty of Medicine and Health Sciences; Universidad de Panamá, Facultad de Medicina; Hospital for Tropical Diseases, London, UK; Institute for Global Health and Infectious Diseases, University of North Carolina at Chapel Hill, Chapel Hill, NC, USA

**Keywords:** COVID-19, sexual behaviour, virtual sex, HIV care, STI testing, Latin America

## Abstract

**Objectives:** To describe perceived changes in sexual behaviours, including virtual sex (sexting and cybersex), and access to HIV/STI testing and care during COVID-19 measures in Panama. **Methods** We conducted an online cross-sectional survey from August 8 to September 12, 2020, among adults (≥18 years) residing in Panama. Participants were recruited through social media. Questions included demographics, access to HIV/STI testing and HIV care and sexual behaviours three months before COVID-19 social distancing measures and during social distancing measures (COVID-19 measures). Logistic regression was used to identify associations between variables and behavioural changes.

**Results:** We recruited 960 participants; 526 (54.8%) identified as cis-women, 366 (38.1%) cis-men, and 68 (7.1%) non-binary or another gender; median age was 28y (IQR:23-37y), 531/957 (55.5%) were of mixed-ethnicity (mixed-Indigenous/European/Afro-descendant ancestry). Before COVID-19 measures, virtual sex was reported by 38.5% (181/470) cis-women, 58.4% (184/315) cis-men and 45.0% (27/60) non-binary participants; during COVID-19 measures, virtual sex increased among 17.2% cis-women, 24.7% cis-men and 8.9% non-binary participants. During COVID-19 measures, 230/800 [28.8%] of participants reported decreased casual sex compared to pre-COVID-19 measures. Compared to pre-COVID-19 measures, decreased casual sex were reported more frequently during COVID-19 measures by cis-men compared to cis-women (39.2% versus 22.9%, urban/rural adjusted odds ratio [AOR]=2.17, 95% confidence interval [CI]:1.57-3.01); and by Afro-descendant compared to mixed-ethnicity participants (40.0% versus 29.8%, AOR=1.78, 95%CI:1.07-2.94). Compared to no change in virtual sex (16.8%), increase in virtual sex (38.5%, AOR=1.78, 95%CI:1.10-2.88); and decreased virtual sex (86.7%, AOR=16.53, 95%CI:7.74-35.27) were associated with decreased casual sex encounters. During COVID-19 measures, HIV/STI testing could not be obtained by 58.0%(58/100) participants who needed a test, and interrupted HIV care was reported by 53.3% (8/15) HIV-positive participants.

**Conclusions:** COVID-19 measures in Panama were associated with a decrease in casual sex among cis-men and Afro-descendant peoples, whilst access to HIV/STI testing and care was seriously disrupted.

## Introduction

HIV and other sexually transmitted infections (STIs) have been on the rise in Panama for the past several years [1], and groups including young adults [2] and Indigenous youth [3] are particularly affected. As of 2019, HIV prevalence nationwide was estimated at 0.6%, belying significant concentrated epidemics among men who have sex with men (MSM) (6.9%) and transwomen (29.6%) populations [1] Correspondingly, STI prevalence is more concentrated among MSM, female sex workers, and adolescents [4-7]. Higher HIV and STI prevalence in Panama is associated with engaging in sexual activity with new and casual partners, due to increased access to connected sexual networks and condomless sex soon after partnership initiation [1].

The COVID-19 pandemic has brought several important changes relevant to sexual health; it also created challenges for in-person data collection. Many population health surveys and other sexual health research activities were initially paused. A small number of published studies from North America and Europe have examined the impact of COVID-19 measures on sexual behaviours [8-10]. However, there is a research gap in Latin America, especially among more isolated rural and Indigenous populations. There is also little information in the region about how COVID-19 measures have affected HIV/STI testing and care services.

Given the relatively high prevalence of HIV and STIs in Panama and the potential for COVID-19 measures to affect both behaviours and access to medical care, the objective of this study was to examine perceived changes in sexual behaviours and access to key sexual health services during COVID-19 measures using an online survey. Based on the association of new and casual partnerships with HIV and STIs, we focused on identifying factors associated with decreased casual sex during COVID-19 measures.

## Methods

This study was an online, cross-sectional survey conducted as part of the first round of International Sexual Health And REproductive Health (I-SHARE), a series of surveys conducted in 34 countries to study sexual and reproductive health during COVID-19 measures [11]. I-SHARE Panama was conducted from August 8 to September 12, 2020 at the end of the strictest COVID-19 lockdown measures (**Figure 1**). Participants reported behaviours from the three-months period before lockdown measures (December 17, 2019 to March 17, 2020) and during the strictest lockdown measures (March 18 to September 12, 2020). The survey was advertised on the website and social media (Facebook and Twitter) of the national public health research institute, Instituto Conmemorativo Gorgas de Estudios de la Salud (ICGES); on social media of non-profit organisations and individuals; and through SMS and direct messages sent to individuals and groups who had previously interacted with partner organizations. Targeted invitation was included in provincial and Comarcal social media platforms to increase participation in these regions to better match census population estimates (**Supplementary Table 1**). Promotion messages asked adults to fill out the survey and/or share the survey link. No IP-address restrictions were included (more than one result can be recorded on the same device) as mobile phones are commonly shared within households and among community members.

**Figure 1:** Timeline of before COVID-19 social distancing measures and COVID-19 social distancing measures in Panama.

### Study design and populations

We used convenience sampling. All adults ages aged ≥18 years who saw the social media/website/messages and lived in Panama were invited to participate.

### Questionnaire creation and study procedures

The questionnaire was collaboratively developed with the I-SHARE consortium [11]. Questions were based on existing survey items and multi-item scales, with some new items developed to address the COVID-19 context [12]. The Panama instrument was translated from the consortium English into Spanish, programmed into OpenDataKit (University of Washington, USA) and pilot-tested with 15 individuals for understanding and acceptability. Participants completed the online questionnaire in 10-30 minutes. Only items associated with skip patterns were obligatory.

Key variables of interest occurring over the <u>three months before</u> and <u>during</u> COVID-19 measures included: sexual intercourse with a casual (new or non-long-term) partner, sexual intercourse with a long-term partner, virtual sex (including ‘sexting’ and ‘cybersex’), use of sexual health services such as HIV/STI testing (“Did COVID-19 measures stop/hinder testing access?”) and HIV care (“Had HIV treatment appointments been cancelled?”). Other influencing variables included: age, sex, gender, number of children, ethnic group, urban/rural residence, household and personal income, sexual orientation, general sexual satisfaction, practice of masturbation, long-term partner variables (cohabitation, tensions, emotional support, cuddling) and condom use with casual and/or long-term partners during the specified time.

### Statistical analyses

We conducted univariable analyses to describe demographic characteristics. We used χ^2^tests to evaluate differences by participants’ sex and other influencing variables and the differences between urban/rural residence; casual sex; and sexual activity with their long-term partner three months before and during COVID-19 measures. All participants with valid data were included; due to non-response on some questions, sample sizes varied.

In addition, we examined factors related to perceived decreased casual sex during COVID-19 measures. We undertook a series of three multivariable analyses comparing participants who reported the frequency of casual sex to have stayed the same over the two periods versus those who reported a decrease during COVID-19 measures, having excluded from analysis individuals who reported an increase in casual sex (n=21). The three models focused on different sets of variables: a) participant socio-demographic variables, b) individual, casual partner variables, c) long-term partners’ behaviours. We first used logistic regression to calculate unadjusted bivariable odds ratios (ORs) and 95% confidence intervals (CIs). Variables associated with the decreased casual sex outcome at p<0.2 level in bivariable analyses were included in multivariable models. As the duration of COVID-19 measures differed between urban and rural regions, we adjusted for residence (urban/rural) in the multivariable analyses. Variables independently associated with decreased casual sex at p<0.1 were included in the final model to provide adjusted odds ratios (AORs) and 95%CIs controlling for participant gender and urban/rural residence. Associations with p<0.05 were considered statistically significant.

### Ethical considerations

We obtained ethical approval from the Comité Nacional de Bioética de Panama (EC-CNBI-2020-06-73), Ghent University (BC-07988) and the University of North Carolina at Chapel Hill for secondary data analysis. Only participants who gave online informed consent by ticking a box could participate. No monetary incentive was provided to participants. The survey did not collect WhatsApp phone numbers, telephone numbers, IP addresses, or any other identifying information.

## Results

In total, participants from 11 out of 12 Panamanian provinces responded to the survey; provincial distribution was similar to the 2020 census projection [13] (**Supplementary Table 1**). Of 960 participants who completed the online questionnaire, 526 (54.8%) identified as cis-women, 366 (38.1%) cis-men and 68 (7.1%) non-binary or of another gender. The median age was 28 years (interquartile range [IQR]: 23-37y). Mixed ethnicity (mixed Indigenous/European/Afro-descendant ancestry) was reported by 55.5% (531/957), Afro-descendent 10.6% (101/957), White 22.2% (212/531), Asian 1.7% (16/957) and Indigenous 10.1% (97/957). Overall, 72.4% (679/938) identified as heterosexual, 7.8% (73/938) as bisexual, 9.6% (90/938) as gay or lesbian, and 10.2% (96/938) as asexual, pansexual, queer, questioning or another orientation (**Table 1**).

**Table 1.** Socio-demographic characteristics of the study population, Panama, 2020.

|  |  | N | % |
| --- | --- | --- | --- |
| **Sex** |  | N=960 |  |
|  | **Female** | 563 | 58.6 |
|  | **Male** | 397 | 41.4 |
| **Gender** |  | N=960 |  |
|  | **Cis-Woman** | 526 | 54.8 |
|  | **Cis-Man** | 366 | 38.1 |
|  | **Non-binary/another gender** | 68 | 7.1 |
| **Age** | **median (28, IQR:23-37)** | N=960 |  |
|  | **18-23** | 280 | 29.2 |
|  | **24-28** | 205 | 21.4 |
|  | **29-37** | 246 | 25.6 |
|  | **38 and above** | 229 | 23.8 |
| **Place of residence** |  | N=958 |  |
|  | **Capital City, large or medium city** | 656 | 68.5 |
|  | **Rural town, community or Comarcal town** | 302 | 31.5 |
| **Number of children** | **Median (0, IQR: 0-1)** | N=960 |  |
|  | **0** | 641 | 66.8 |
|  | **1-2** | 250 | 26.0 |
|  | **3 and more** | 69 | 7.2 |
| **Highest completed level of education** |  | N=957 |  |
|  | **Secondary or less** | 127 | 13.3 |
|  | **Some university** | 289 | 30.2 |
|  | **Post-secondary (university completed)** | 541 | 56.5 |
| **Ethnic group** |  | N=957 |  |
|  | **Mixed\*** | 531 | 55.5 |
|  | **Afro-descendent** | 101 | 10.6 |
|  | **White** | 212 | 22.2 |
|  | **Asian** | 16 | 1.7 |
|  | **Indigenous** | 97 | 10.1 |
| **Successful following of COVID-19 measures** |  | N=957 |  |
|  | **Not at all or only some** | 75 | 7.8 |
|  | **Mostly or strictly follow them** | 882 | 92.2 |
| **Have you been in strict confinement due to COVID-19** |  | N=955 |  |
|  | **No** | 817 | 85.5 |
|  | **Yes** | 138 | 14.5 |
| **SARS-CoV-2 test** |  | N=956 |  |
|  | **Never** | 775 | 81.1 |
|  | **Yes, and positive at least once** | 26 | 2.7 |
|  | **Yes, and always been negative** | 155 | 16.2 |
| **Including yourself, how many people do you live with** | **median 3 people (IQR2-4)** | N=943 |  |
|  | **0** | 33 | 3.5 |
|  | **1** | 118 | 12.5 |
|  | **2** | 233 | 24.7 |
|  | **3 to 4** | 439 | 46.6 |
|  | **5 or more** | 120 | 12.7 |
| **Change in employment** |  | N=935 |  |
|  | **No change in what I do or going to work** | 183 | 19.6 |
|  | **Work from home part-time or fulltime** | 448 | 47.9 |
|  | **lost work or without work before and during COVID-19 measures** | 304 | 32.5 |
| **Household income since COVID started** |  | N=926 |  |
| | **US\$0 a \$499 monthly** | 273 | 29.5 |
| | **US\$500 a \$999 monthly** | 176 | 19.0 |
| | **US\$1000 a \$2000 monthly** | 206 | 22.3 |
| | **US\$2001-\$5000 monthly** | 195 | 21.1 |
| | **US\$5001 and higher monthly** | 76 | 8.2 |
| **Changes in household economic situation since COVID started** |  | N=953 |  |
|  | **Household economics got worse** | 483 | 50.7 |
|  | **No changes in household economics** | 445 | 46.7 |
|  | **Household economics got better** | 25 | 2.6 |
| **Weekly Alcohol use during COVID-19 social distancing** |  | N=932 |  |
|  | **Decreased** | 427 | 45.8 |
|  | **The same** | 349 | 37.4 |
|  | **Increased** | 156 | 16.7 |
| **Sexual orientation** |  | N=938 |  |
|  | **Heterosexual** | 679 | 72.4 |
|  | **Bisexual** | 73 | 7.8 |
|  | **Gay or Lesbian** | 90 | 9.6 |
|  | **Asexual, pansexual, queer, questioning, another gender** | 96 | 10.2 |

\*mixed ethnicity is mixed Indigenous/European/Afro-descendant/Asian ancestry

### Sexual behaviours

Previous sexual experience was reported by 88.8% (852/960) of participants. Before COVID-19 measures, casual sex among sexually experienced participants was reported by 18.2% (85/466) of cis-women, 32.45 (101/312) of cis-men and 23.7% (14/59) of non-binary participants (**Table 2, Panel A)**. Of those who reported casual sex, always using a condom in such encounters was reported by 50.6% (43/85) of cis-women, 61.2% (63/103) cis-men and 53.9% (7/13) non-binary participants (**Supplementary Table 2**). Of all participants, compared to before COVID-19 measures, during COVID-19 measures, 68.6% (549/800) of respondents reported no change, 28.8% (230/800) reported a decrease, and 2.6% (21/800) reported an increase in casual sex (**Table 2, Panel A**).

**Table 2.** Sexual behaviours during COVID-19 social distancing measures in Panama, Panama, 2020.

|  |  | All participants |  | Cis-women |  | Cis-men |  | Non-binary/another gender |  |
| --- | --- | --- | --- | --- | --- | --- | --- | --- | --- |
|  |  | N | % | N | % | N | % | N | % |
| **Panel A: Individual and casual partner behaviours** |  |  |  |  |  |  |  |  |  |
| **Sexual Satisfaction** |  | N=833 |  | N=463 |  | N=311 |  | N=59 |  |
|  | Decreased | 406 | 48.7 | 243 | 52.5 | 133 | 42.8 | 30 | 50.8 |
|  | The same | 211 | 25.3 | 115 | 24.8 | 81 | 26.1 | 15 | 25.4 |
|  | Increased | 216 | 25.9 | 105 | 22.7 | 97 | 31.9 | 14 | 23.7 |
| **Sexual Problems\*** |  | N=486 |  | N=301 |  | N=156 |  | N=29 |  |
|  | Decreased | 240 | 49.4 | 140 | 46.5 | 83 | 53.2 | 17 | 58.6 |
|  | The same | 141 | 29.0 | 89 | 29.6 | 44 | 28.2 | 8 | 27.6 |
|  | Increased | 105 | 21.6 | 72 | 23.9 | 29 | 18.6 | 4 | 13.8 |
| **Masturbated** |  | N=817 |  | N=455 |  | N=307 |  | N=55 |  |
|  | Decreased | 181 | 22.1 | 120 | 26.4 | 46 | 15.0 | 15 | 27.3 |
|  | The same | 410 | 50.2 | 246 | 54.1 | 132 | 43.0 | 32 | 58.2 |
|  | Increased | 226 | 27.7 | 89 | 19.6 | 129 | 42.0 | 8 | 14.6 |
| **Pornography use** |  | N=804 |  | N=442 |  | N=306 |  | N=56 |  |
|  | Decreased | 141 | 17.5 | 78 | 17.7 | 52 | 17.0 | 11 | 19.6 |
|  | The same | 483 | 60.1 | 306 | 69.2 | 138 | 45.1 | 39 | 69.6 |
|  | Increased | 180 | 22.4 | 58 | 13.1 | 116 | 37.9 | 6 | 10.7 |
| **Virtual sex\*\*** |  | N=800 |  | N=441 |  | N=303 |  | N=56 |  |
|  | Decreased | 94 | 11.8 | 36 | 8.2 | 45 | 14.9 | 13 | 23.2 |
|  | The same | 547 | 68.4 | 329 | 74.6 | 180 | 59.4 | 38 | 67.9 |
|  | Increased | 159 | 19.9 | 76 | 17.2 | 78 | 24.7 | 5 | 8.9 |
| **Casual sex encounters** |  | N=800 |  | N=441 |  | N=304 |  | N=55 |  |
|  | Decreased | 230 | 28.8 | 99 | 22.4 | 115 | 37.8 | 16 | 29.1 |
|  | The same | 549 | 68.6 | 333 | 75.5 | 178 | 58.5 | 38 | 69.1 |
|  | Increased | 21 | 2.6 | 9 | 2.0 | 11 | 3.6 | 1 | 1.8 |
| **Condom use with a casual partner** |  | N=198 |  | N=81 |  | N=102 |  | N=15 |  |
|  | Decreased | 35 | 17.7 | 12 | 14.8 | 19 | 18.6 | 4 | 26.7 |
|  | The same | 131 | 66.2 | 59 | 72.8 | 63 | 61.8 | 9 | 60.0 |
|  | Increased | 32 | 16.2 | 10 | 12.3 | 20 | 19.6 | 2 | 13.3 |
| **Panel B: Long-term relationship behaviours** |  |  |  |  |  |  |  |  |  |
| **Long-term partnership tensions\*\*\*** |  | N=495 |  | N=308 |  | N=159 |  | N=28 |  |
|  | Decreased | 169 | 34.1 | 107 | 34.7 | 51 | 32.1 | 11 | 39.3 |
|  | The same | 158 | 31.9 | 102 | 33.1 | 47 | 29.6 | 9 | 32.1 |
|  | Increased | 168 | 33.9 | 99 | 32.1 | 61 | 38.4 | 8 | 28.6 |
| **Sex with a long-term partner** |  | N=491 |  | N=305 |  | N=157 |  | N=29 |  |
|  | Decreased | 249 | 50.7 | 153 | 50.2 | 82 | 52.2 | 14 | 48.3 |
|  | The same | 178 | 36.2 | 108 | 35.4 | 61 | 38.8 | 9 | 31.0 |
|  | Increased | 64 | 13.0 | 44 | 14.4 | 14 | 8.9 | 6 | 20.7 |
| **Condom use with a long-term partner** |  | N=479 |  | N=298 |  | N=154 |  | N=27 |  |
|  | Decreased | 77 | 16.1 | 47 | 15.8 | 24 | 15.6 | 6 | 22.2 |
|  | The same | 386 | 80.6 | 244 | 81.9 | 122 | 79.2 | 20 | 74.1 |
|  | Increased | 16 | 3.3 | 7 | 2.3 | 8 | 5.2 | 1 | 3.7 |

\*Sexual problems are individual or partners issues including erectile dysfunction or inhibited desire, orgasm
\*\*virtual-sex is a composite variable of cybersex use and/or sexting use
\*\*\*Long-term partner relationship tensions **Fight with long-term partner**

Of participants with a long-term partner, sex with the long-term partner and casual sex encounter at least monthly before COVID-19 measures was reported by 18.2% (80/440) of cis-women, 33.5% (93/278) cis-men, and 25.5% (14/55) non-binary participants.

Overall, before COVID-19 measures, 47.1% (394/837) of participants reported being sexually satisfied (**Supplementary Table 2**). Of those not satisfied before COVID-19 measures, 33.9% (150/442) reported increased satisfaction during COVID-19. During COVID-19 measures, 21.6% (105/486) reported an increase in sexual problems (individual or partner’s erectile dysfunction or inhibited desire, orgasm) and 49.4% (240/486) reported a decrease in sexual problems **Table 2)**.

Before COVID-19 measures, 46.4% (392/845) participants reported using virtual sex at least once a month, including sexting 44.1% (369/837) and cybersex 20.4% (172/842). Use of virtual sex had increased during COVID-19 measures for 19.9% (159/800) of participants, decreased for 11.8% (94/800), and remained unchanged for 68.4% (547/800) (**Table 2, Panel A**).

### Long-term partner relationship and sexual behaviours

Overall, 66.4% (637/960) participants reported having a long-term relationship before COVID-19 measures; 504 (79.1%) of whom reported to still be in their long-term relationship during COVID-19 measures. Sexual intercourse at least monthly with their long-term partner was reported by 92.7% (332/358) cis-women, 82.5% (160/194) cis-men and 90.9% (40/44) non-binary participants. Decreases in sex with their long-term partner during COVID-19 measures was reported by 50.2% (153/305) cis-women, 52.2% (82/157) cis-men, and 48.3% (14/29) non-binary participants (**Table 2, Panel B**).

### Access to HIV/STI testing and HIV care services

Overall, 45.6% (58/823) participants reported that condoms were more difficult to find during COVID-19 measures. This percentage did not differ between urban and rural areas (44.2% [273/299] urban compared to 49.5% [101/204] rural, p=0.38).

Of the 10.4% (100/960) who reported they needed an STI or HIV test, 58.0% (58/100) reported they could not receive it due to the COVID-19 measures. This percentage was higher among urban areas, however the difference was not significant (62.0% [44/71] urban compared with 48.3% [14/29] rural, p=0.20).

Few (15/960, 1.6%) participants reported to be living with HIV, 8 of whom reported to have had an HIV care appointment cancelled or postponed due to COVID-19 measures. Thirteen of the 15 respondents living with HIV were in urban areas, and all participants living with HIV reported worrying about ART shortages.

### Factors related to decreased casual sex during COVID-19

#### Participant characteristics

After adjusting for urban/rural residence, cis-men were more likely to report decreases in casual sex during COVID-19 measures (39.2%) compared to cis-women (22.9%) (AOR=2.16, 95%CI:1.58-2.95) (**Table 3, Panel A)**.

**Table 3:**
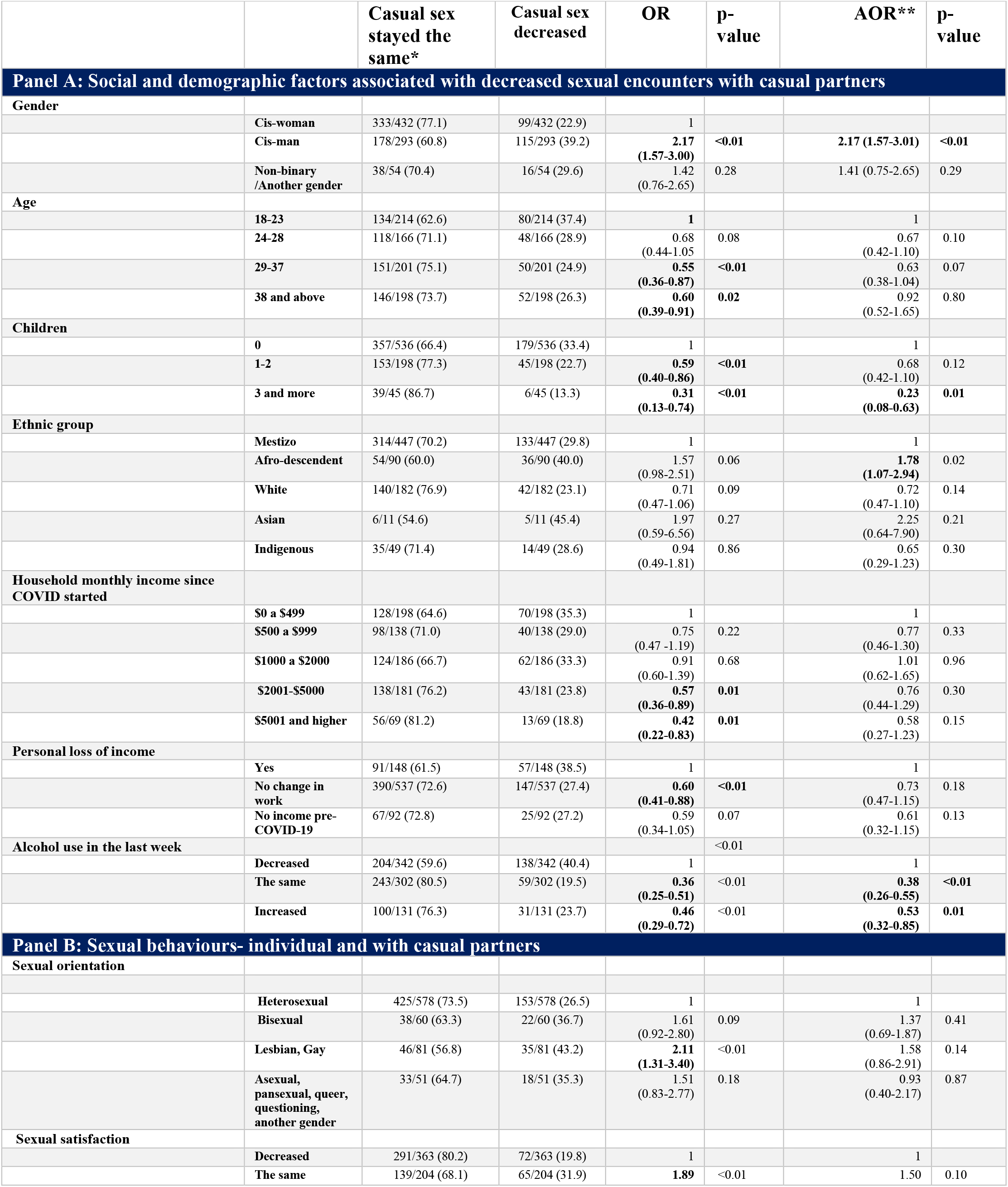
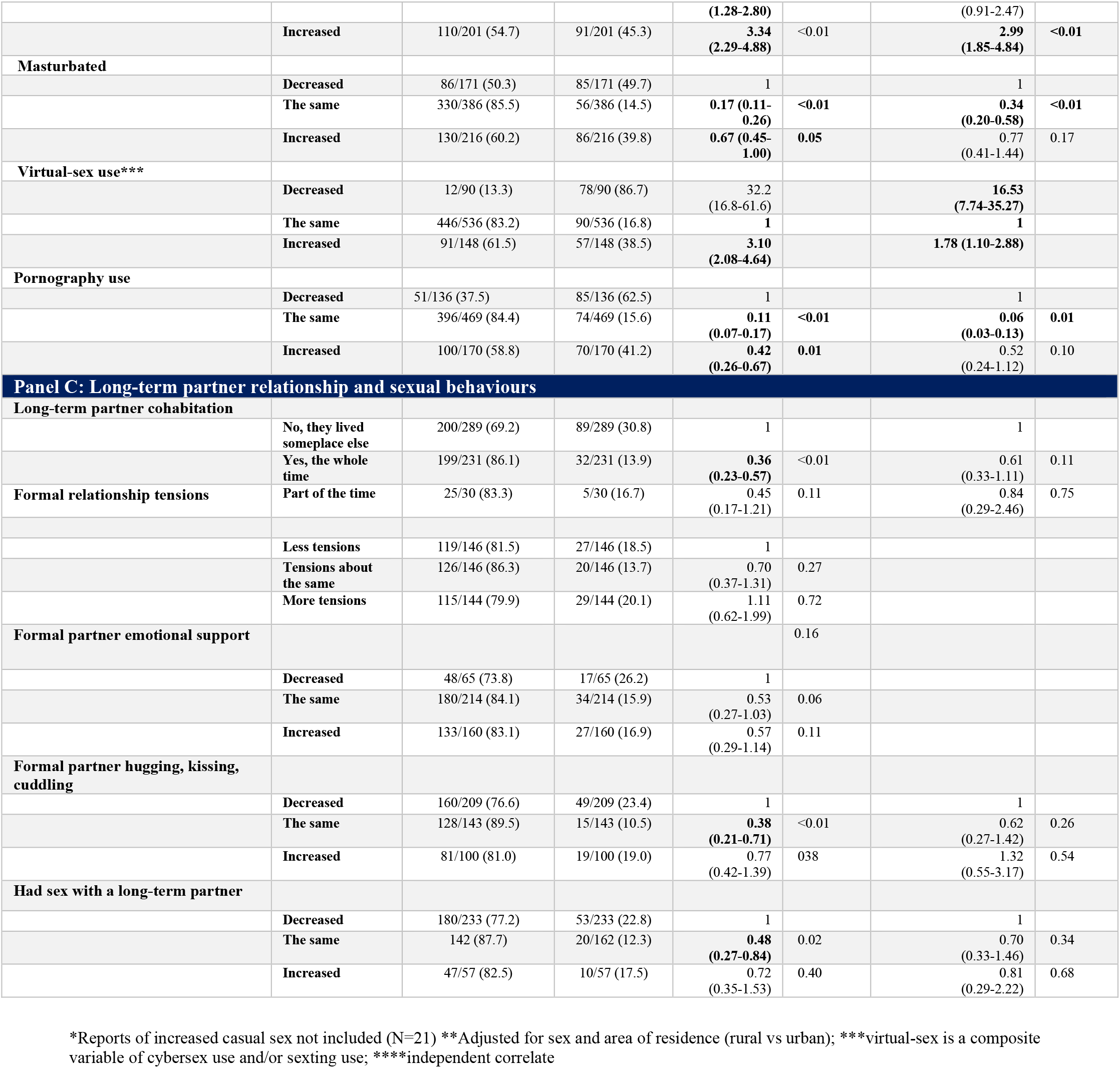
Demographic, social and sexual factors associated with a reported decrease in casual sex partners during COVID-19 measures in Panama, 2020.

After adjusting for participant gender and urban/rural residence, several factors were associated with decreased casual sex during COVID-19 measures. Individuals of Afro-descendant ethnicity reported a larger decrease in sex with casual partners (40.0% compared to 29.8% among mixed ethnicity, AOR=1.78, 95%CI:1.07-2.94). There was weak association between sex orientation and decreased sex with casual partners (43.2% among gay or lesbian participants versus 26.5% of heterosexual participants, AOR=1.66, 95%CI:0.88-3.14); (**Table 3, Panel B**).

Participants who reported the same or increased levels of alcohol use during COVID-19 measures were less likely to report decreased casual sex compared with those who decreased their alcohol use during COVID-19 measures (19.5% and 23.7% versus 40.4%, AOR=0.38, 95%CI:0.26-0.55 and AOR=0.53, 95%CI:0.32-0.85, respectively).

#### Individual sexual behaviours and virtual sex use

After adjusting for participant gender and urban/rural residence, an increase in sexual satisfaction was associated with a decrease in casual sex: 45.3% of those reporting an increase in sexual satisfaction also reported decreased casual sex compared to 19.8% among those reporting decreased sexual satisfaction (AOR=3.34, 95%CI:2.29-4.88).

Perceived changes in virtual sex during COVID-19 measures were also associated with decrease in casual sex in adjusted models. Compared to no change in virtual sex (16.8%), an increase in virtual sex (38.5%) was associated with decreased casual sex (AOR=1.78,95%CI:1.10-2.88) and decreased virtual sex (86.7%,AOR=16.53, 95%CI:7.74-35.27) were associated with decreased casual sex.

## Discussion

This study examined perceived changes in sexual behaviours, use of virtual sex, and access to HIV/STI testing and HIV care during the implementation of COVID-19 measures in Panama. Our results among a diverse convenience sample of urban and rural dwellers across 11/12 provinces expand the literature about sexual behaviours during COVID-19 measures in Latin America. We found that overall sexual activity may have decreased among some individuals. Casual sex, widely practised by 18.2% of cis-women, 32.4% of cis-men and 23.7% of nonbinary participants pre-COVID, decreased for 22.9% of cis-women, 39.1% of cis-men and 29.6% of nonbinary participants. On the other hand, virtual sex, also widely practised by 20 to 40% of respondents pre-COVID-19, increased for 20% of respondents. Finally, participants reported COVID-19 measures interrupted access to condoms, HIV/STI testing and, worryingly, to HIV care for those who needed the services.

A large proportion of participants reported decreased sexual activity during COVID-19 measures, findings that differ from the Latvian I-SHARE study, which found most individuals did not have a change sexual frequency during COVID-19 measures [14]. Half of Panama participants reported decreased sex with a long-term partner. This may have been due to extended periods together and increased time with children or other housemates [8, 15]. Sex with a casual partner decreased among more than one-quarter of individuals. Sex with casual partners has also shown to have decreased in the United States and Australia early in the pandemic [8, 10, 16]. A decrease in casual sex partners may provide a unique opportunity for a reduction in behavioural risk, but further research is needed.

Nearly half of our participants reported engaging in virtual sex before COVID-19 measures; 20% used cybersex, and 40% used sexting at least monthly, including in very rural provinces. Twenty percent of participants reported increased virtual sex during COVID-19 measures. Before COVID-19 measures, cybersex use in Sweden (32%) was found to be slightly higher than what was found in Panama [17]. Some researchers and media outlets had hypothesised that virtual sex might increase during COVID-19 due to fewer in-person sexual encounters. However, the first analyses from North America did not demonstrate this [9, 10]. Interestingly, we found that participants who reported to have either increased or decreased virtual sex use were more likely to report a decrease in casual sex, compared to those who did not change their virtual sex use.

Our findings indicate that virtual sex use in Panama may serve both as a substitute and a preamble to in-person sex. A pre-COVID-19 meta-analysis found positive correlations between sexting, number of sex partners and condomless sex [18]. As virtual sex practices emerge as normalized, sex-positive tools of sexual behaviour in Panama, privacy and potential extorsion warnings should be addressed within the applications themselves. Additionally, community-wide campaigns could educate on privacy laws and recommend use of encrypted applications [19].

The HIV epidemic is concentrated among specific populations, particularly MSM and transwomen; STI prevalence is high among adolescents and unregistered female sex workers [4-7]. Regular testing of HIV/STI in these populations is considered to aid in controlling transmission. Interruption of HIV/STI services may lead to decreased diagnoses and treatment, thereby increasing continued transmission and increase in sequelae. During COVID-19 measures in Panama, access to key services was interrupted, with over 50% of those needing HIV/STI testing not getting it. This is supported by an overall decrease of 71% of new HIV diagnoses reported by the Panama government during that period [20]. A study in Australia also found a substantial decrease in HIV tests during 2020 [21]. HIV testing services elsewhere, including

Latin America, have been significantly interrupted during the global pandemic [22-26]. Respondents also reported disrupted access to STI and HIV prevention (condoms) and HIV care commodities. Such difficulties in accessing HIV/STI testing and care in Panama during the COVID-19 measures may have been related to limited transportation, testing facility closures or the *covidization* of health services. Panama does not have policies supporting HIV self-testing and STI sample self-collection, as can be found elsewhere globally. Therefore, our findings suggest the need for patient self-testing approaches [27, 28] that would help maintain continuity of services during national medical crises.

Our study has several limitations. First, online questionnaires are likely to suffer selection bias as they are only able to include participants who have seen the announcement and are both motivated and able to use online tools. However, our study used best practices for the conduct of online research during COVID-19 [29]. These recommendations include using an online panel for the study design, implementing the survey with partner organizations, designing the survey for the end-user experience, and having a prespecified analysis plan [29]. We found that our sample had a similar structure to the 2020 census data in terms of ethnicity, urban/rural residence, and province of residence (**Supplementary Table 1**), except that our sample had more female participants compared to census data. This finding is common in health surveys that examine sexual and reproductive health topics [30]. Thirdly, casual sexual encounters may be underreported as over half of our participants were in long-term partnerships; additionally, memory bias may have enhanced or impaired recall of behaviours from before and during COVID-19 measures. Fourthly, comparisons between sexual orientation and gender groups should be interpreted with caution given the relatively smaller sample of non-cisgender respondents. Fifthly, this paper focused on casual sexual encounters and sexual behaviours in general. Other related sexual health topics, including intimate partner violence, access to reproductive health services and mental health will be reported elsewhere. Sixthly, as this is a single behavioural cross-sectional study, and the capacity for collecting biological samples was limited during this time, therefore there should be caution when making any casual inferences from the data. Lastly, this analysis is unable to correlate behaviour with biological outcomes, therefore we do not know the impact of behaviour changes on HIV or STI rates.

Our study has implications for STI and HIV research and policy. Our data suggest the need and usefulness of more rigorous behavioural research during national medical crises and pandemics that have the capacity to disturb normal services. While we were able to recruit a convenience sample during the pandemic, national panels or other methods can be used to obtain less biased observations [29]. These data will be important as lockdown conditions are relaxed and reinforced. From a policy perspective, our data underline the importance of maintaining the continuity of HIV/STI testing and care services even during emergency responses.

## Conclusions

Our findings add to the sexual behaviour literature in Panama during COVID-19. We found a decrease in sexual activity among some individuals for casual encounters, paralleled with a rise in the use of virtual sex. STI and HIV prevention and care services were significantly disrupted during COVID-19 measures, suggesting the need for decentralized services.

## Data Availability

Data are available under reasonable request to the corresponding author.

## Acknowledgements

We are thankful to all the participants of this study, and to all who shared the study links on social media. JMP is a distinguished member of the National Research System that is supported by the National Secretariat of Science, Technology and Innovation. This study was conducted under the umbrella of the I-SHARE study (International Sexual Health and REproductive Health), which examines the impact of the COVID-19 crisis on sexual and reproductive health in diverse low-income, middle-income, and high-income countries. The full list of consortium members and their roles can be found here (https://ishare.web.unc.edu).

## Key Messages

- During Panama’s COVID-19 social distancing measures, we found a decrease in sexual activity among some individuals, especially casual sex encounters among cis-men and participants of Afro-descendent ethnicity
- Virtual sex (sexting and cybersex) use was common before COVID-19 social distancing measures. Perceived changes in this practice were associated with a decrease in casual sex.
- Condom access, STI and HIV testing, and HIV care were seriously interrupted during COVID-19 social distancing measures in Panama.

## Contributorship statement

Authors AG, JTE, KM, CPE, MM, JK, GCT, MdA, AM and JT contributed to the conception of the international and national versions of the I-SHARE project and survey instrument. Data collection was performed by AG, JK, GCT, MdA, AM and JMP. Data curation and analysis was performed by AG, JTE, JT, PM. AG drafted the manuscript, all authors contributed to critical revisions and final approval. AG is responsible for the overall content as a guarantor.

---

**Supplementary Table 1:** Comparison of ISHARE-Panama sample to Census projected 2020 population 18 years and older.

|  | Census projected population per province or Comarca [1] |  | ISHARE-Panama Sample |  |
| --- | --- | --- | --- | --- |
|  | Raw population 18 years and older | % of the population | Total number recruited | % of the total sample |
| *Province/ Indigenous Comarca* |  |  |  |  |
| *Total population* | 2937150 |  | 960 |  |
| *Bocas* | 102885 | 3.5% | 15 | 1.6% |
| *Chiriquí* | 311118 | 10.6% | 82 | 8.5% |
| *Coclé* | 183592 | 6.2% | 46 | 4.8% |
| *Colón* | 191773 | 6.5% | 52 | 5.4% |
| *Comarca Ngäbe-Buglé* | 144423 | 4.9% | 58 | 6.0% |
| *Herrera* | 85178 | 2.9% | 19 | 2.0% |
| *Los Santos* | 70372 | 2.4% | 14 | 1.5% |
| *Panama and Panama Oeste* | 1611313 | 54.9% | 643 | 67.0% |
| *Veraguas* | 166561 | 5.7% | 26 | 2.7% |
| *Comarca Guna* | 26286 | 0.9% | 0 | 0.0% |
| *Darien* | 36279 | 1.2% | 2 | 0.2% |
| *Comarca Embera-Wounaan* | 7370 | 0.2% | 1 | 0.1% |

**Supplementary Table 2.**
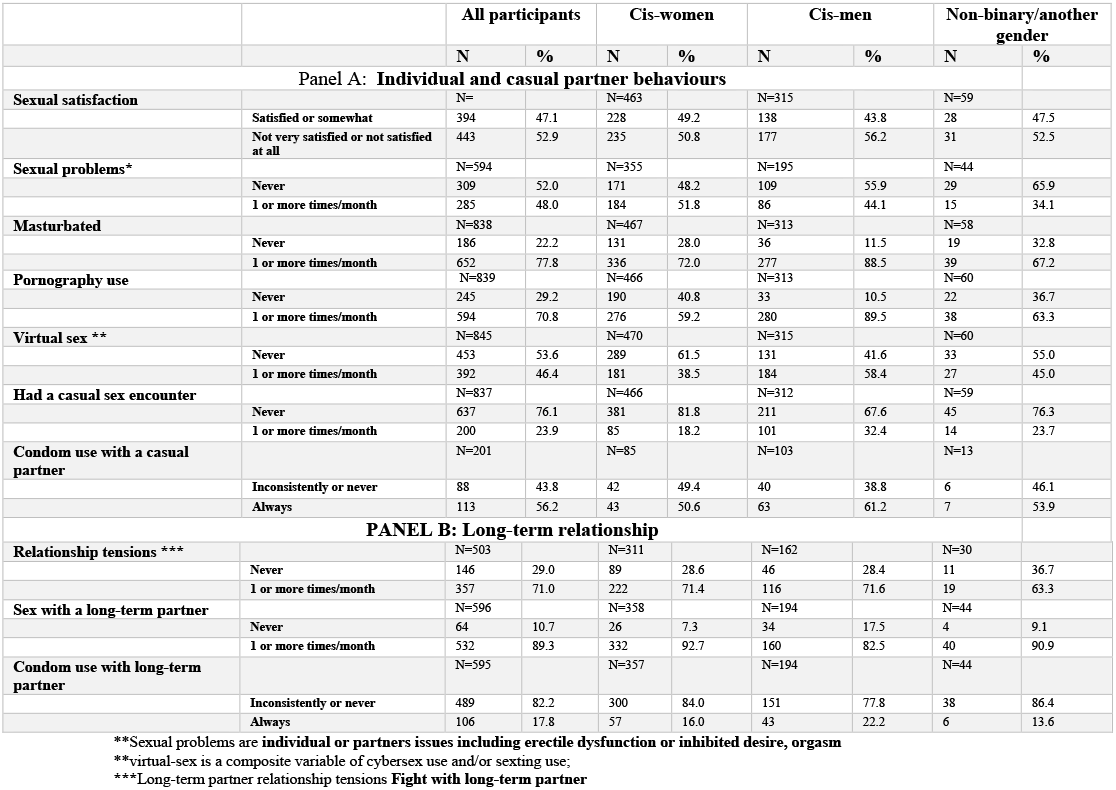
Sexual behaviours three months before COVID-19 social distancing measures in anama, Panama, 2020.

