## Supplementary files for "How did COVID-19 measures impact sexual behaviour and access to HIV/STI services in Panama? Results from a national cross-sectional online survey" (from another version of this preprint: version 1, 2021-02-05)

**Supplementary Table 1:** Comparison of ISHARE-Panama sample to Census projected 2020 population 18 years and older

|  | Census projected population per province or Comarca [1] | | ISHARE-Panama  Sample | |
| --- | --- | --- | --- | --- |
| **Province/**  **Indigenous Comarca** | **Raw population 18 years and older** | **% of the population** | **Total number recruited** | **% of the total sample** |
| **Total population** | 2937150 |  | 960 |  |
| **Bocas** | 102885 | 3.5% | 15 | 1.6% |
| **Chiriquí** | 311118 | 10.6% | 82 | 8.5% |
| **Coclé** | 183592 | 6.2% | 46 | 4.8% |
| **Colón** | 191773 | 6.5% | 52 | 5.4% |
| **Comarca Ngäbe-Buglé** | 144423 | 4.9% | 58 | 6.0% |
| **Herrera** | 85178 | 2.9% | 19 | 2.0% |
| **Los Santos** | 70372 | 2.4% | 14 | 1.5% |
| **Panama and Panama Oeste** | 1611313 | 54.9% | 643 | 67.0% |
| **Veraguas** | 166561 | 5.7% | 26 | 2.7% |
| **Comarca Guna** | 26286 | 0.9% | 0 | 0.0% |
| **Darien** | 36279 | 1.2% | 2 | 0.2% |
| **Comarca Embera-Wounaan** | 7370 | 0.2% | 1 | 0.1% |

Reference to Supplementary table 1:

1.	Instituto Nacional de Estadistica y Censo. Boletin 15. Estimaciones y proyecciones de la poblacion en la republica, provincia, comarca indigena por distrito segun sexo y edad 2010-2020. 2010.

**Supplementary Table 2. Sexual behaviours three months before COVID-19 social distancing measures in Panama, Panama, 2020.**

|  | |  | | All participants | | | Cis-women | | | | Cis-men | | | | | Non-binary/another gender | | |
| --- | --- | --- | --- | --- | --- | --- | --- | --- | --- | --- | --- | --- | --- | --- | --- | --- | --- | --- |
|  | |  | | **N** | | **%** | **N** | | **%** | | **N** | | | **%** | | **N** | | **%** |
| Panel A: Individual and casual partner behaviours | | | | | | | | | | | | | | | | | |  |
| Sexual satisfaction | |  | | N= | |  | N=463 | |  | | N=315 | | |  | | N=59 | |  |
|  | | **Satisfied or somewhat** | | 394 | | 47.1 | 228 | | 49.2 | | 138 | | | 43.8 | | 28 | | 47.5 |
|  | | **Not very satisfied or not satisfied at all** | | 443 | | 52.9 | 235 | | 50.8 | | 177 | | | 56.2 | | 31 | | 52.5 |
| Sexual problems* | |  | | N=594 | |  | N=355 | |  | | N=195 | | |  | | N=44 | |  |
|  | | **Never** | | 309 | | 52.0 | 171 | | 48.2 | | 109 | | | 55.9 | | 29 | | 65.9 |
|  | | **1 or more times/month** | | 285 | | 48.0 | 184 | | 51.8 | | 86 | | | 44.1 | | 15 | | 34.1 |
| Masturbated | |  | | N=838 | |  | N=467 | |  | | N=313 | | |  | | N=58 | |  |
|  | | **Never** | | 186 | | 22.2 | 131 | | 28.0 | | 36 | | | 11.5 | | 19 | | 32.8 |
|  | | **1 or more times/month** | | 652 | | 77.8 | 336 | | 72.0 | | 277 | | | 88.5 | | 39 | | 67.2 |
| Pornography use | |  | | N=839 | |  | N=466 | |  | | N=313 | | |  | | N=60 | |  |
|  | | **Never** | | 245 | | 29.2 | 190 | | 40.8 | | 33 | | | 10.5 | | 22 | | 36.7 |
|  | | **1 or more times/month** | | 594 | | 70.8 | 276 | | 59.2 | | 280 | | | 89.5 | | 38 | | 63.3 |
| Virtual sex ** | |  | | N=845 | |  | N=470 | |  | | N=315 | | |  | | N=60 | |  |
|  | | **Never** | | 453 | | 53.6 | 289 | | 61.5 | | 131 | | | 41.6 | | 33 | | 55.0 |
|  | | **1 or more times/month** | | 392 | | 46.4 | 181 | | 38.5 | | 184 | | | 58.4 | | 27 | | 45.0 |
| Had a casual sex encounter | |  | | N=837 | |  | N=466 | |  | | N=312 | | |  | | N=59 | |  |
|  | | **Never** | | 637 | | 76.1 | 381 | | 81.8 | | 211 | | | 67.6 | | 45 | | 76.3 |
|  | | **1 or more times/month** | | 200 | | 23.9 | 85 | | 18.2 | | 101 | | | 32.4 | | 14 | | 23.7 |
| Condom use with a casual partner | |  | | N=201 | |  | N=85 | |  | | N=103 | | |  | | N=13 | |  |
|  | | **Inconsistently or never** | | 88 | | 43.8 | 42 | | 49.4 | | 40 | | | 38.8 | | 6 | | 46.1 |
|  | | **Always** | | 113 | | 56.2 | 43 | | 50.6 | | 63 | | | 61.2 | | 7 | | 53.9 |
| PANEL B: Long-term relationship | | | | | | | | | | | | | | | | | |  |
| Relationship tensions *** |  | | N=503 | |  | | | N=311 | |  | | N=162 |  | | N=30 | |  | |
|  | **Never** | | 146 | | 29.0 | | | 89 | | 28.6 | | 46 | 28.4 | | 11 | | 36.7 | |
|  | **1 or more times/month** | | 357 | | 71.0 | | | 222 | | 71.4 | | 116 | 71.6 | | 19 | | 63.3 | |
| Sex with a long-term partner |  | | N=596 | |  | | | N=358 | |  | | N=194 |  | | N=44 | |  | |
|  | **Never** | | 64 | | 10.7 | | | 26 | | 7.3 | | 34 | 17.5 | | 4 | | 9.1 | |
|  | **1 or more times/month** | | 532 | | 89.3 | | | 332 | | 92.7 | | 160 | 82.5 | | 40 | | 90.9 | |
| Condom use with long-term partner |  | | N=595 | |  | | | N=357 | |  | | N=194 |  | | N=44 | |  | |
|  | **Inconsistently or never** | | 489 | | 82.2 | | | 300 | | 84.0 | | 151 | 77.8 | | 38 | | 86.4 | |
|  | **Always** | | 106 | | 17.8 | | | 57 | | 16.0 | | 43 | 22.2 | | 6 | | 13.6 | |

**Sexual problems are **individual or partners issues including erectile dysfunction or inhibited desire, orgasm**

**virtual-sex is a composite variable of cybersex use and/or sexting use;

*******Long-term partner relationship tensions **Fight with long-term partner**
